# Concurrent virtual reality and transcranial alternating current stimulation for social cognition and neural activity in schizophrenia: A proof-of-concept study

**DOI:** 10.1101/2025.05.09.25327343

**Authors:** Kirsten Gainsford, Bernadette M. Fitzgibbon, Aron T. Hill, Paul B. Fitzgerald, Caroline T. Gurvich, Kate E. Hoy

**Affiliations:** Department of Psychiatry, School of Translational Medicine, School of Medicine, Nursing and Health Sciences, Monash University, Vic, Australia; School of Medicine and Psychology, Australian National University, Canberra, ACT, Australia; Cognitive Neuroscience Unit, School of Psychology, Deakin University, Melbourne, Vic, Australia; The Bionics Institute of Australia, 384-388 Albert St, East Melbourne, Vic, Australia

## Abstract

Social cognition, including theory of mind (ToM), is impaired in people with schizophrenia which can significantly impact daily functioning. Current interventions for social cognitive impairment are often time consuming and have limited ecological validity. Combining emerging technologies such as virtual reality (VR) and transcranial alternating current stimulation (tACS) may help address these limitations. The current study applied theta tACS to the right temporoparietal junction (rTPJ) during VR social cognition training in 15 participants with schizophrenia. Neurophysiological (event-related potentials and spectral power) and behavioural outcome measures (ToM task performance) were assessed. Participants underwent two experimental sessions. One session involved VR with concurrent active theta tACS (5Hz frequency) and the other consisted of VR with concurrent sham theta tACS. Resting state electroencephalography (EEG) and ToM tasks with concurrent EEG were measured pre- and post- VR-tACS. Order of stimulation condition was randomised, and stimulation and assessments were all double-blinded. We found ToM task response time improved after VR, regardless of tACS condition. While only VR and active tACS, but not sham, resulted in a widespread increase in resting state theta power. This is the first study to combine VR and tACS in a psychiatric population to address social cognition and provides initial evidence to support the feasibility and efficacy of a combined VR-tACS protocol in schizophrenia. Implications for future research are discussed.

## 1. Introduction

Social cognitive skills are critical for understanding and interpreting the social world (Arioli, Crespi, & Canessa, 2018). Social cognitive difficulties are well documented in people with schizophrenia (Green, 2016; Green, Horan, & Lee, 2019). In particular, theory of mind (ToM: the ability to take others’ perspectives, predicting and interpreting their intentions and beliefs) has consistently been shown to be impaired in people with schizophrenia (Green et al., 2019) and to contribute to poor functioning (Fett et al., 2011; Halverson et al., 2019; Healey, Penn, Perkins, Woods, & Addington, 2013). Difficulties in ToM are present throughout the lifetime of the disorder with people at the early stages of schizophrenia performing worse at ToM tasks than controls but better than people with chronic schizophrenia, suggesting a worsening of ToM abilities over time (Canty, Neumann, & Shum, 2017).

There are a number of social cognitive interventions used in schizophrenia, focusing on either broad (i.e., multiple social cognitive domains) or specific skill improvement (for a review see Tan, Lee, & Lee, 2018). While these interventions have been shown to improve performance on specific standardised measures of social cognition, they have been shown not to generalise well beyond these, with little impact on day-to-day functioning (Yeo, Yoon, Lee, Kurtz, & Choi, 2022). This may be due to these interventions taking place clinical settings and/or consisting of computer-based training; settings that are not representative of the real-world environments where social cognitive skills are needed (d’Arma et al., 2021; Yeo et al., 2022). Virtual reality (VR) social cognition training is a more ecologically valid approach (Hoşgelen, Güneri, Erdeniz, & Alptekin, 2024), and it is becoming more commonly used in schizophrenia with evidence showing higher effect sizes for VR social cognition training compared to traditional methods (Shen, Liu, Wu, Lin, & Wang, 2022). VR can also be combined with technologies such as non-invasive brain stimulation (NIBS) to further enhance the effects of therapeutic interventions (Cassani, Novak, Falk, & Oliveira, 2020). For example, techniques such as transcranial alternating current stimulation (tACS) can be used in conjunction with VR to enhance activity in specific cortical regions. This is achieved by using an electrical current administered at specific frequencies to entrain target brain activity (Antal & Paulus, 2013).

Of particular relevance, the right temporoparietal junction (rTPJ) is a key brain region subserving ToM and is part of social brain networks involved in widespread activation during ToM processes (Igelström & Graziano, 2017). It has been shown to be less active and less functionally connected to other social brain areas in people with schizophrenia (Bitsch, Berger, Nagels, Falkenberg, & Straube, 2021; Penner et al., 2018) and reduced rTPJ activity has also been directly associated with reduced ToM abilities in schizophrenia (Lee, Quintana, Nori, & Green, 2011). Theta oscillations in the rTPJ have been found to be important for both functioning within the rTPJ and more widespread communication between the rTPJ and broader social brain networks (Seymour, Wang, Rippon, & Kessler, 2018). Theta oscillations are also reduced in schizophrenia when engaging in cognitive tasks (Ryman et al., 2018; Uhlhaas & Singer, 2010). Taken together this would suggest enhancing theta activity in the rTPJ may help to improve brain activity underlying social cognition. This can be achieved using theta-tACS. No studies have investigated the impact of tACS on ToM at the rTPJ in schizophrenia. However, in populations with similar social cognitive difficulties such as autism spectrum disorder (ASD), a recent study showed that four sessions of transcranial direct current stimulation (tDCS) to the rTPJ improved performance in false belief and self-other judgement tasks testing ToM compared to sham tDCS (Padrón et al., 2022). Combining tACS or non-invasive brain stimulation (NIBS) with VR training could also enhance these effects.

VR-NIBS treatment is a growing field of interest, which, to date, has been investigating stroke rehabilitation (motor skills recovery), multiple sclerosis (balance and fatigue), neuropathic pain (analgesic effects of adding NIBS), cerebral palsy (mobility and balance) and phobias or post-traumatic stress disorder (emotion regulation and anxiety reduction; Cassani et al., 2020). VR-NIBS studies have shown the potential to be more effective for rehabilitation than either intervention alone. For example, a combined VR-tDCS protocol to improve upper extremity functioning after stroke, showed scores on fine motor skills tasks were significantly higher when participants received VR-tDCS compared to either VR or tDCS alone (Lee & Chun, 2014). Thus far, no research has investigated the effects of combing VR and NIBS in schizophrenia. We recently conducted a proof-of-concept study in a healthy control sample to investigate feasibility of VR-tACS for social cognition with promising results, including behavioural improvements in ToM task accuracy after participants received active theta tACS during VR social cognition training as well as an effect of VR training on resting state theta power (Gainsford et al., 2025a). The current study will, for the first time, investigate the effects of concurrent VR-tACS on social cognition in a schizophrenia sample.

The current study aimed to investigate the feasibility and efficacy of combining VR and theta frequency tACS to improve ToM in participants with a diagnosis of schizophrenia using neurophysiological and behavioural outcome measures. In this proof-of-concept pilot study we hypothesised that ToM task accuracy and response time would show a significant improvement after concurrent VR and active theta tACS (i.e., VR+tACS) to the rTPJ as compared to concurrent VR and sham theta tACS (i.e., VR+Sham). We also hypothesised that resting state theta power would be significantly increased following VR+tACS as compared to VR+Sham. Finally, we also examined the impact of VR-tACS on event-related potentials (ERP) associated with ToM processes. Specifically, the late positive component (LPC) which is initiated in centro-parietal brain areas between 300-700ms after a ToM stimulus is presented and the temporoparietal 450 (TP450) which arises 325-525ms after a ToM stimulus in the rTPJ region, peaking at 450ms (Libsack et al., 2022; Vistoli et al., 2015).

## 2. Methods

### 2.1 Design

This was a double-blind, randomised, sham-controlled, within-subjects pilot study. Participants each attended three visits. A baseline visit, where a cognitive and social cognitive task battery were completed. Data from the baseline visit have been published previously and are not the focus of this paper (Gainsford et al., 2025b). Participants then attended two experimental sessions, which were conducted at least 72 hours apart (**Figure 1**). Participants were randomly allocated to active or sham tACS for experimental session 1 and they received the opposite protocol in the second experimental session. At all times during both experimental sessions, participants wore an EEG cap containing both EEG and tACS electrodes. Participants wore this combined EEG-tACS cap before, during, and after VR-tACS. EEG cords were stowed in a backpack while the participant completed VR-tACS. Detailed tACS blinding and randomisation information is outlined in Section 2.6 below.

**Figure 1:**
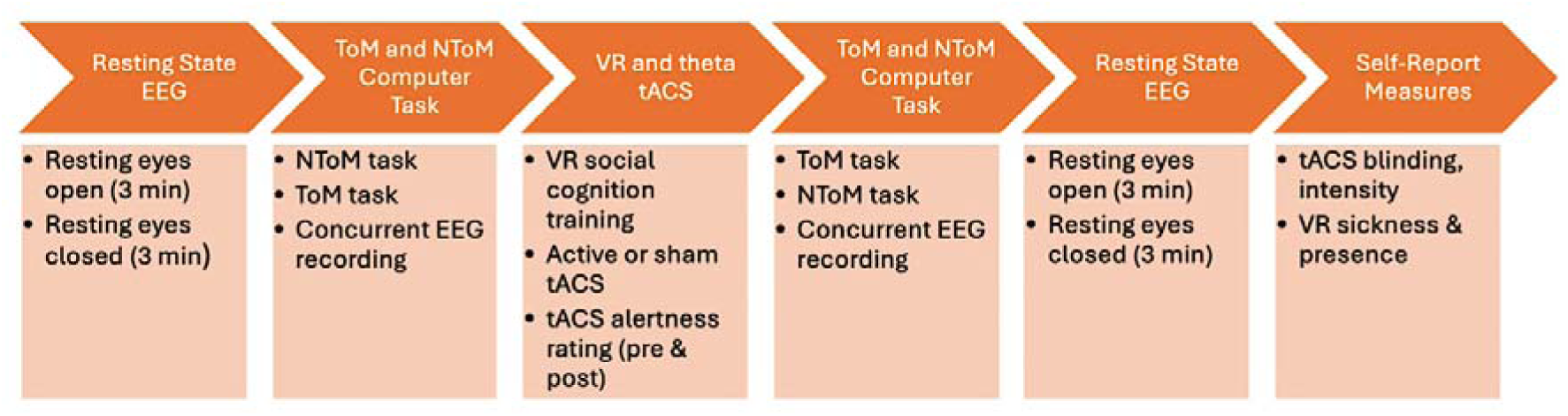
Protocol of experimental sessions 1 and 2. First, resting state EEG with eyes open and eyes closed were conducted. Then non-theory of mind (NToM) task followed by the theory of mind (ToM) task. To carry out the VR-tACS, tACS impedance was first conducted, VR head mounted display (HMD) put on the participant and VR-tACS completed. After VR-tACS, computer tasks were done again using the versions of each task not yet completed with ToM tasks being first. Finally, resting state EEG recordings were repeated and then VR and tACS experience self-report questionnaires done.

### 2.2 Participants

Fifteen participants (9F, 6M, M = 43.27) form the final pilot sample including nine participants with schizophrenia and six with schizoaffective disorder (see **Table 1** for descriptives). Participant medications and treatments are outlined in **Table 2**. Participants on medications were on stable doses for at least four weeks prior to entering the trial. Participants were excluded if they were not stable on their medications or symptoms were not stable. Participants were also excluded if they had alcohol or substance use disorders, had experienced significant traumatic brain injury, history of seizures or epilepsy or other neurological disorders as well as any unstable medical or psychiatric conditions. Schizophrenia diagnosis was confirmed using the mini international neuropsychiatric interview (MINI; Sheehan, 2016) and the positive and negative schizophrenia scale (PANSS; Kay, Opler, & Lindenmayer, 1989) was conducted to determine symptom severity.

**Table 1:** Descriptives of schizophrenia sample.

|  | Measure | M(SD) | Range |
| --- | --- | --- | --- |
| PANSS |  |  |  |
|  | Positive | 15.533(5.630) | 7-24 |
|  | Negative | 15.267(4.511) | 10-24 |
|  | General | 31.867(6.081) | 22-40 |
|  | Total | 62.667(13.189) | 40-78 |
| Education<br>(Yrs) |  | 14.80(2.077) | 10-19 |
| Years since<br>diagnosis |  | 17.20(7.063) | 7-34 |
| PANSS = Positive and Negative Syndrome Scale |  |  |  |

**Table 2:**
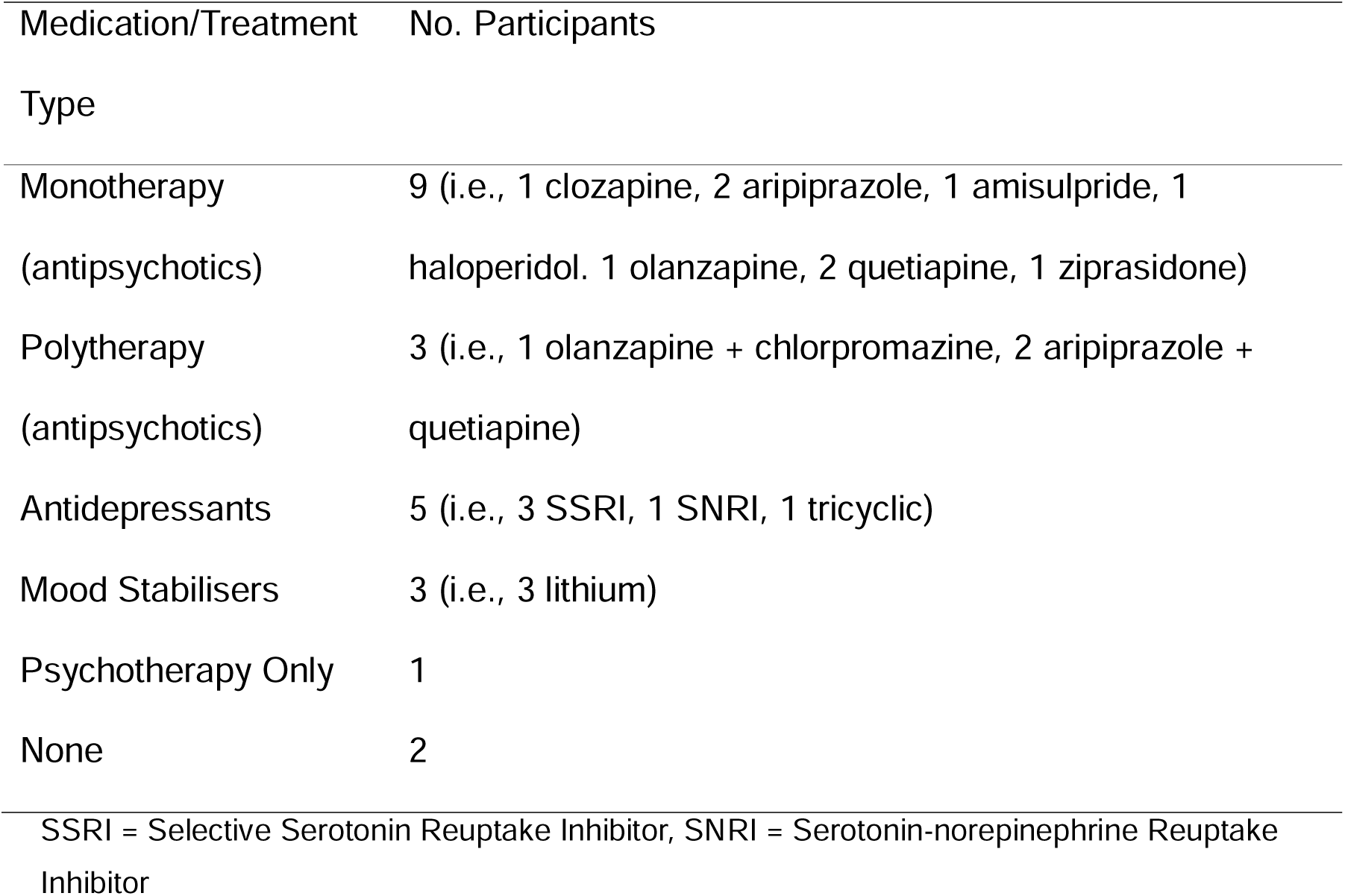
Participant medications and treatments.

The study took place at the Epworth Centre for Innovation in Mental Health (ECIMH) between 2021 and 2022, and then the Monash Alfred Psychiatry Research Centre (MAPrc) between 2022 and 2023. All participants had their data collected at a single site, and sites were set up to be consistent for data collection purposes. Participants were primarily recruited via referral from our participant database consisting of people who previously took part in our research and consented to being contacted for future studies. Other recruitment avenues were referral from community support services (e.g., hospitals, housing care, social support services, GP or psychiatry clinics) and social media and recruitment platforms (e.g., HealthMatch).

All participants provided written informed consent. Ethics approval was obtained from the Monash Health Human Research Ethics Committee and governance obtained through Monash University, Alfred Health and Epworth HealthCare.

### 2.3 EEG

A 50-channel EEG montage (see **Figure 4**) utilizing electrodes from the 10-05 system (ActiCap, Brain Products, Gilching, Germany) was used to record data via the Brain Vision Recorder software (Brain Products, Gilching, Germany). Ag/AgCl electrodes were connected to a BrainAmp amplifier (Brain Products, Gilching, Germany). The custom EEG setup was designed to accommodate both tACS electrodes and a VR headset. The electrodes included in the setup were: AF3, AFz (ground), AF4, F7, F5, F3, F1, Fz, F2, F4, F6, F8, FC5, FC3, FC1, FCz, FC2, FC4, FC6, T7, C5, FCC6h, CCP5h, C3, C1, Cz, C2, C4, FCC6h, CCP6h, T8, TP7, CP5, CP3, CP1, CPz (reference), CP2, CP4, CP6, TP8, P7, P5, P3, P1, Pz, P6, P8, PO3, POz, PO4, O1, Oz, and O2. Impedances were routinely monitored and kept below 10 kΩ. Data was recorded at a sampling rate of 1000Hz, with a high-pass filter set to 0.01Hz and a low-pass filter at 200Hz.

The EEG cap remained on the participant’s head throughout the stimulation sessions. During the VR-tACS protocol, the electrode cables were unplugged and placed in a bag on the participant’s back. After completing VR-tACS, the VR headset was removed. Depending on the testing location, the lab equipment was returned to its original position, or the participant was moved back to the EEG lab. The EEG system was then reconnected, impedance checked, and the ToM behavioural task was initiated. Participants began the ToM task approximately five minutes after completing the VR-tACS procedure, with the order of task versions counterbalanced.

#### 2.3.1 EEG Resting State

EEG recordings were conducted for three minutes each resting with eyes open (REO) and resting eyes closed (REC), both before and after participants completed VR-tACS within each session. Participants were seated approximately one meter away from the computer screen and were asked to remain as still as possible while recordings were taken. For the REO, they were instructed to focus on a fixation cross at the centre of the screen, and for REC, they were instructed to relax with their eyes shut.

### 2.4 Behavioural Task

Participants completed the attribution of intentions Theory of Mind (ToM) task in each session both before and after the VR-tACS protocol. This task is an accepted measure for assessing ToM (Eddy, 2019). Simultaneous EEG was recorded each time the task was performed. The task was developed using Inquist 6 (Millisecond, Seattle WA, USA, 2020) and based on the work of Vistoli et al. (2015), employing the same set of stimuli. However, since the protocol included multiple sessions, the stimuli were split into different task versions.

The set of stimuli consisted of 82 comic strips, divided into three categories. Each comic included four images, with both congruent and incongruent endings. The intention attribution task required participants to use ToM to interpret the comic’s content (i.e., the ToM task). The physical causality with objects (PCOb) and physical causality with characters (PCCh) tasks did not require ToM and were grouped together to form the non-Theory of Mind (NToM) task.

There were two versions of the ToM task (ToM A, ToM B) and two versions of the NToM task (NToM A, NToM B). Each ToM version consisted of 24 trials, split evenly between congruent and incongruent endings. Each NToM version had 48 trials, with 24 congruent and 24 incongruent endings. The remaining ten comics were used for two shorter practice tests. Task versions A and B each contained either the congruent or incongruent ending of the comics, with an online random number generator used to assign comics to task versions. Comics were not repeated within each version, and the order of comics within each task was randomised. Each task version was presented once per session, with task order counterbalanced.

Participants sat approximately one meter from the computer screen while performing the tasks. For the ToM task, they were instructed to assess whether the fourth image in each comic “makes sense for the person’s goals.” In the NToM task, they were asked to determine whether the fourth image “makes sense as the ending of the story.” Participants responded by pressing a green button on a button box if the fourth image made sense, or a red button if it did not make sense. The timing of the comic sequence is shown in **Figure 2**. The fourth image remained on the screen until the participant responded or for 4000ms, whichever was first. To help participants know when to respond, a red square outlined the fourth image of each comic (**Figure 2**). The number of correct responses and response times were recorded.

**Figure 2:**
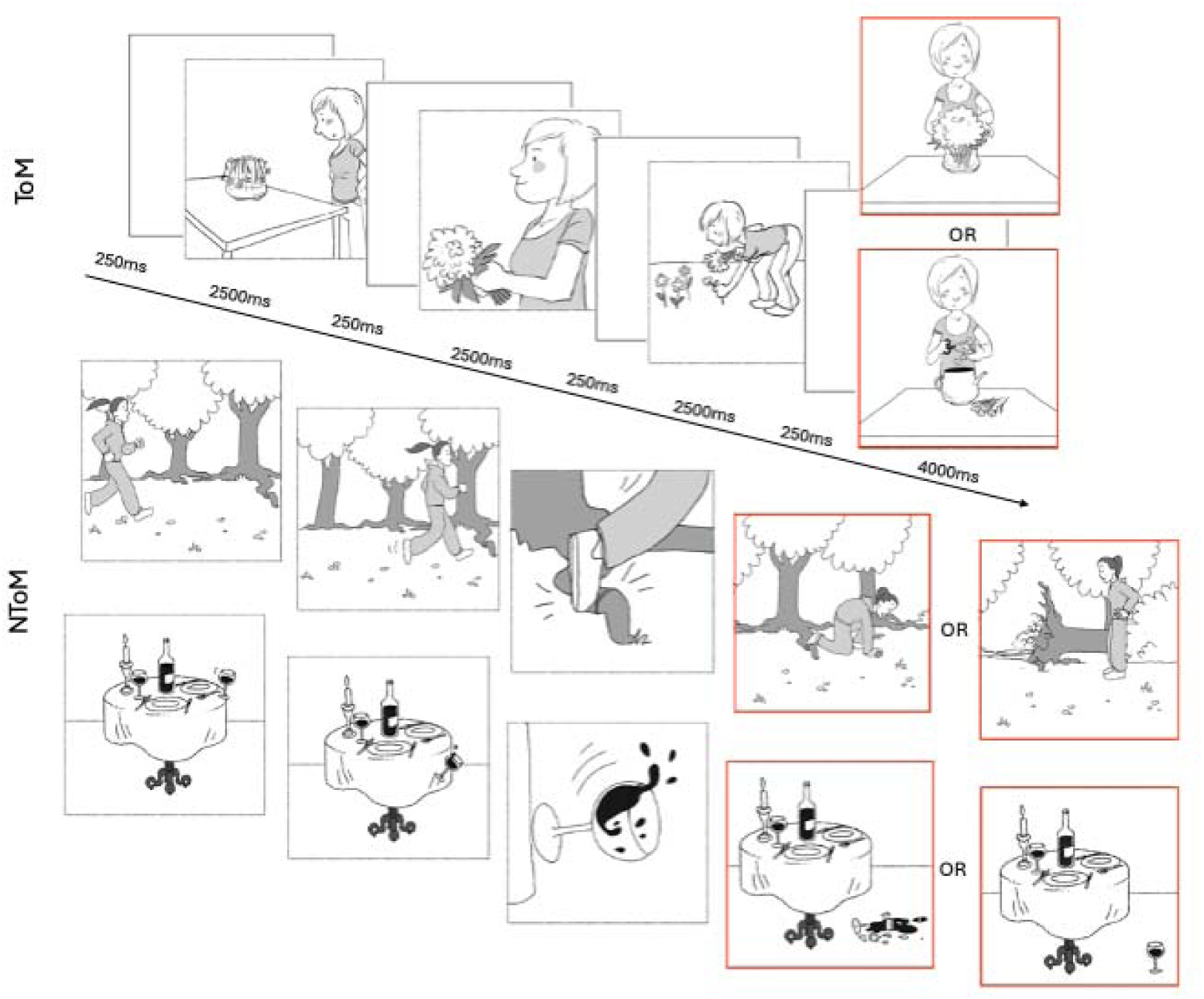
Attribution of intentions ToM task. The first is a ToM example with congruent or incongruent ending options and NToM examples (PCCh and PCOb) with alternate endings below. EEG was recorded while participants completed this task (extracted from Gainsford et al., 2025b).

### 2.5 VR

Participants wore a Meta Quest 2 head-mounted display (HMD; Meta, 2020) to complete the VR tasks using hand-held controllers. At both research locations, the lab equipment was arranged to allow participants to move freely and safely while performing the VR task. They navigated through a virtual shopping mall and restaurant (see **Figure 3**) in a custom-designed VR program (SoCog, SensiLab, Monash University, Melbourne Australia). During the task, participants interacted with virtual avatars to answer questions interpreting their actions or emotions and chose appropriate responses from a set of options. The program provided feedback on the accuracy of their responses, supporting learning. The VR task was completed by participants once during both the active and sham tACS sessions, for a total of two times.

**Figure 3:**
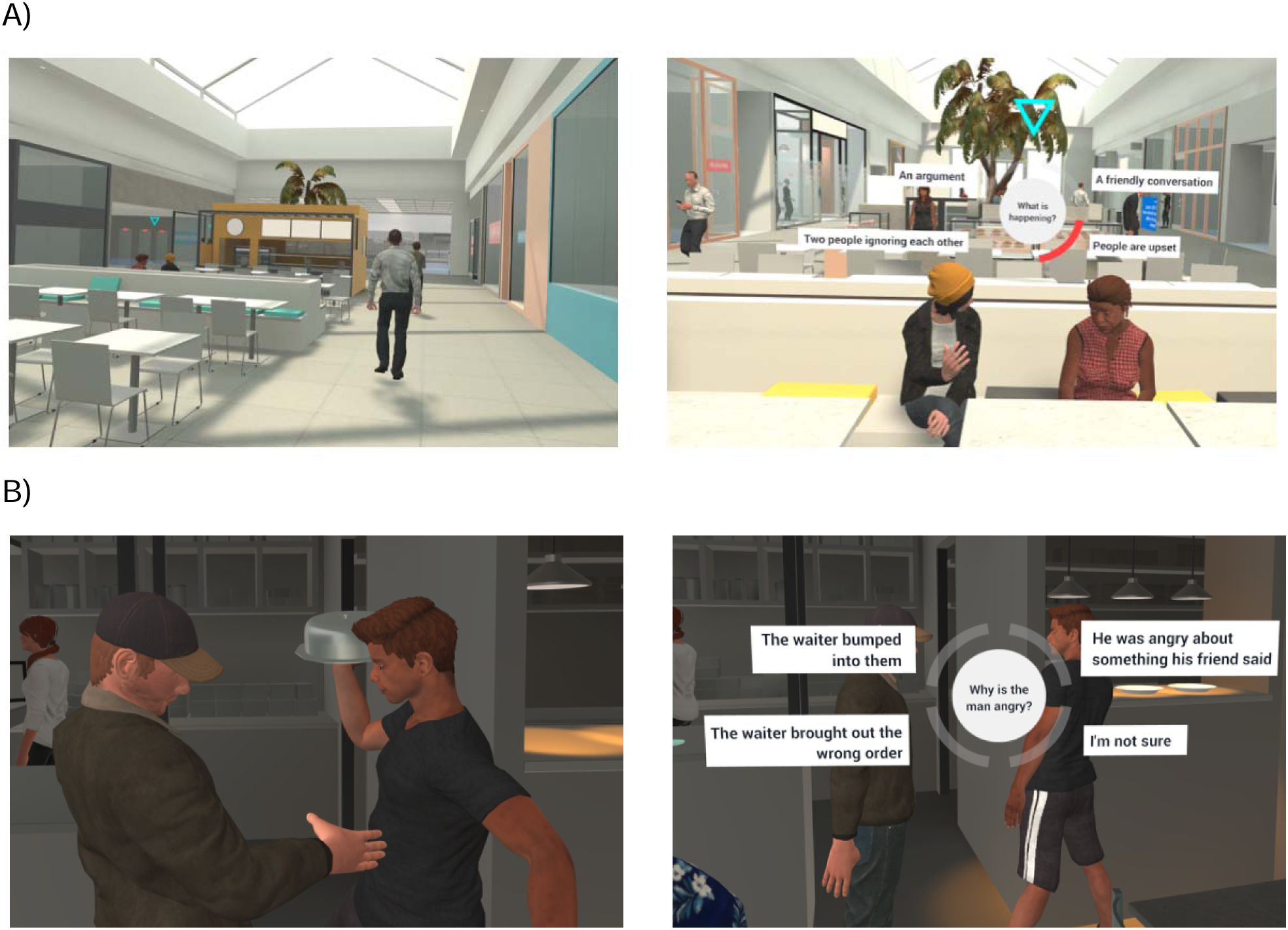
Examples of virtual environments participants encountered in VR social cognition training. A) Participants explored a shopping mall while encountering tasks such as the one on the right where they had to determine what type of conversation is happening based on the avatar’s movements, behaviours and social cues. B) Participants had a meal at a café and were required to infer others’ thoughts when they encountered a conflict.

### 2.6 tACS

The active tACS protocol involved a 30-second ramp-up phase at the start, followed by 15 minutes of theta frequency stimulation at 5Hz, and concluded with a 30-second ramp-down, totalling 16 minutes. In contrast, the sham protocol consisted of a 30-second ramp-up immediately followed by a 30-second ramp-down, commonly used in research due to it simulating the sensation of tACS without inducing neurophysiological effects. E-field current modelling was performed using SimNIBS software (Version 3.2, Saturnino, Madsen, & Thielscher, 2020). A montage was developed to maximise current flow over the cortical regions associated with the rTPJ using the software (MNI coordinates: 50, −53, 35; see **Figure 4**). Prior to the main study, a small pilot was conducted of three different stimulation intensities using the selected montage (C4, C6, P2, P4). Users were asked about degree of scalp sensations and discomfort to determine the most appropriate stimulation intensity to use. The total current intensity was set to 1.75mA and tACS was administered using the StarStim^®^ wireless hybrid EEG/tCS 8-channel system (Neuroelectrics, Barcelona, Spain) along with the NIC2 program (Neuroelectrics, Barcelona, Spain). Pi stim Ag/AgCl electrodes (3.14 cm²) were filled with saline gel, and the tACS impedance was maintained below 10 kΩ.

**Figure 4:**
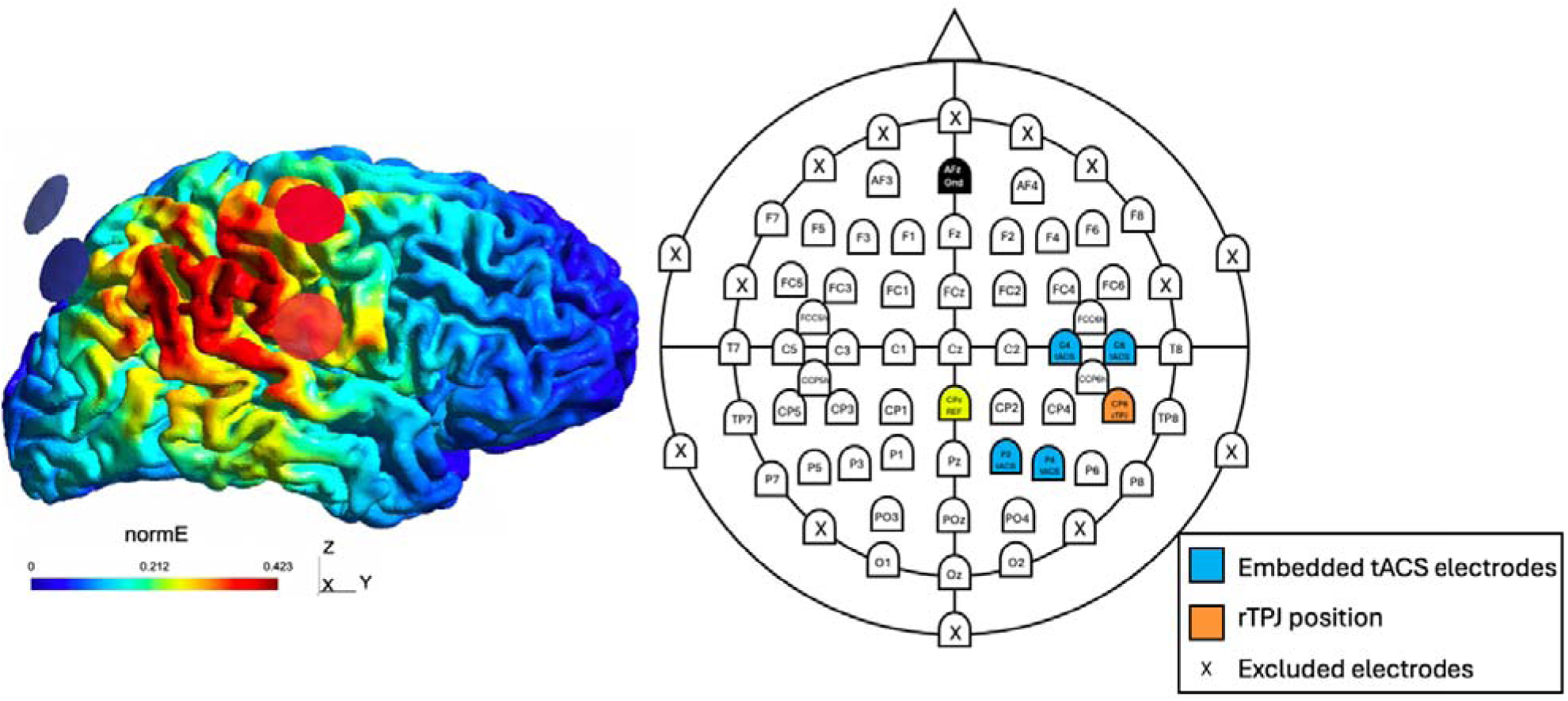
tACS E-field modelling (left) and custom EEG, tACS montage (right). The left side of the figure shows the current modelling conducted with SimNIBS to determine electrode placement for theta tACS. Electrodes were embedded in the EEG cap at C4, C6, P2 and P4. Red circles represent stimulation electrodes (C4, C6) and blue circles represent return electrodes (P2, P4). The image on the right shows the EEG montage with embedded tACS electrodes (blue) and rTPJ highlighted (orange). The “X’s” indicate where electrodes were removed for the comfort of the VR headset (frontally) and to improve efficiency with EEG set up.

Both the active and sham protocols were programmed into NIC2, with an unblinded team member (uninvolved in data collection or analysis) assigning a code (A or B) to each protocol. The double-blind mode was activated using a password known only to the unblinded researcher. Participants were randomly assigned to receive either active or sham tACS in their first stimulation session, with the opposite protocol applied in the second stimulation session. Randomisation was conducted using Microsoft Excel, which assigned letters A or B to numbers from 1 to 26. Each participant completed one active and one sham session of tACS. Once set up in the virtual environment, participants began the VR tasks after the tACS ramp-up period (30 seconds) was complete. They performed the VR tasks while receiving tACS. If the VR task finished before tACS ended, participants were seated and placed in a calm virtual environment until the conclusion of stimulation.

### 2.7 tACS Blinding, Intensity and Alertness

In each experimental session, participants were asked to guess whether they were receiving active or sham stimulation and to indicate their confidence level on a scale from 0 (“complete guess”) to 10 (“absolutely certain”). Participants were also asked to rate their level of alertness on a scale of 0 (“very drowsy”) to 10 (“very alert”) before and after stimulation. Following the stimulation, they rated the intensity of the sensation of tACS on a scale from 0 (“no sensation”) to 10 (“extremely strong”) and provided feedback on any other sensations or side effects they experienced during the stimulation.

### 2.8 VR Sickness and Presence

After each VR-tACS session, participants completed the VR Sickness Questionnaire (VRSQ; Kim, Park, Choi, & Choe, 2018). They rated the extent to which they experienced side effects during the VR task on a scale from 0 (“not at all”) to 3 (“very”). Sub-scores for oculomotor symptoms (general discomfort, fatigue, eyestrain, difficulty focusing) and disorientation symptoms (headache, fullness of head, blurred vision, dizziness, vertigo) were calculated, along with a total score. At the end of each session, participants also completed the Presence Questionnaire (PQ; Witmer & Singer, 1998) to evaluate their level of immersion in the VR experience. Responses were provided on a Likert scale from 1 to 7, with 7 representing the highest. Sub-scores were derived by summing the total scores for the following items: “realism”, “possibility to act”, “quality of interface” (reverse-scored), “possibility to examine”, “self-evaluation” of performance, and “sounds”.

### 2.9 Analysis

#### 2.9.1 Task Response Accuracy and Response Time

Response accuracy was assessed by averaging the percentage of correct responses of each participant. To calculate mean response times, the time point at which participants responded to the fourth picture of each comic was extracted and then averaged. If participants made a response error (i.e., responded on the button box before the presentation of the fourth picture in the comic), the trial was excluded. Pairwise t-tests were conducted using JASP (Version 0.17.3, JASP Team, Amsterdam, Netherlands 2023) to determine the statistical differences in accuracy and response time for pre versus post active and sham VR-tACS. When investigating the difference between stimulation conditions (i.e., VR+tACS vs VR+Sham), difference scores were calculated by subtracting pre-from post-stimulation accuracy scores and response times for both active and sham. Then, paired samples t-tests were used to statistically compare the data.

#### 2.9.2 EEG Analysis

All EEG data were pre-processed in MATLAB (2024a, Mathworks, 2024) using a fully automated EEG pre-processing pipeline, namely, RELAX v1.1.5 (Bailey, Biabani, et al., 2023; Bailey, Hill, et al., 2023) with the EEGLAB toolbox (Delorme & Makeig, 2004). Please see Gainsford et al. (2025a) for a more detailed description of data cleaning processes. Epochs were created after data were cleaned and for resting state analysis, were separated into 3 second epochs (−1.5 to 1.5 sec) with no overlap (additional information in supplementary materials). Epochs were defined around the third image of each comic for ERP analysis as this was the point of the story where ToM neural processes would likely be activated (Vistoli et al., 2015) and were 1.3 seconds in total (−300ms to 1000ms, additional information in supplementary materials). FieldTrip (Oostenveld, Fries, Maris, & Schoffelen, 2011) was used to average each participants’ epochs from pre- and post-tACS for resting data and ERP data to create grand average files.

##### 2.9.2.1 Resting State EEG Cluster Analysis

To calculate spectral power, a fast Fourier transform (FFT) of 1-45Hz at 0.5Hz intervals was performed on the pre-processed data and epoched to convert it into the frequency domain. Grand average files were calculated for each condition (REO and REC, pre and post, active and sham). A whole-brain data driven approach was used to conduct resting state cluster-based permutation analyses, to broadly identify areas of cortical activation at rest (5000 permutations). Customised MATLAB scripts were used to analyse change in in theta power (4-8Hz) at rest. Within-subjects cluster analyses were conducted for pre versus post VR+tACS and VR+Sham. Resting data were statistically compared using two-tailed within subjects (dependent samples) t-tests (α = 0.05, two-tailed) to determine effect of stimulation on resting state theta power, post VR+tACS and post VR+Sham.

##### 2.9.2.2 Resting State EEG ROI Analysis

To analyse theta band activity (4-8Hz) at rest, electrodes representing the rTPJ (CP4, CP6, TP8, P6, P8) were selected. Mean amplitudes at these electrodes were extracted for each participant and inputted into JASP. Paired samples t-tests were used to compare resting state theta power post-compared to pre-VR-tACS within VR+tACS and VR+Sham sessions. To compare active and sham conditions, post-VR-tACS, mean amplitudes were subtracted from pre- to create difference scores for active and sham sessions. The data was then compared in JASP using paired samples t-tests.

##### 2.9.2.3 Event-Related Potential Analysis

Two ERPs were investigated that relate to ToM processes. First, the temporoparietal 450 (TP450), peaking at approximately 450ms, was examined. With the analysis timepoint of interest classified as 325ms to 525ms. To conduct ROI analyses, we investigated changes in peak amplitude at the CP6 electrode (representing the rTPJ) and a broader group of electrodes around the rTPJ (CP4, CP6, TP8, P6, P8). The second type of ERP examined was the late positive component (LPC), occurring centro-parietally between 300ms and 700ms. Changes in peak amplitude were analysed at Pz (parietal midline) alone and Pz and Cz together (centro-parietal midline; taken as an average of the signal across the two electrodes). This was determined based on the literature. To determine change in theta power post VR-tACS compared to pre within VR+tACS and VR+Sham sessions, the mean amplitude across the aforementioned electrodes were extracted and statistically compared in a series of paired-samples t-tests in JASP. To compare active and sham conditions, post-VR and tACS, mean amplitudes were subtracted from pre- to create difference scores for VR+tACS and VR+Sham sessions prior to conducting paired samples t-tests. As an additional ‘whole brain’ data-driven approach, cluster-based analyses were conducted to explore which cortical regions activated during task performance. Two-tailed dependent samples t-tests were conducted to identify significant electrode clusters pre versus post VR-tACS within each session and post versus post VR+tACS and VR+Sham (α = 0.05, two-tailed; Maris & Oostenveld, 2007).

#### 2.9.3 Correlations

Using MATLAB and FieldTrip, significant electrode clusters from resting state cluster-based permutation analyses were extracted. For each participant and each condition (i.e., REO and REC pre and post VR+tACS and VR+Sham), the mean amplitude of theta power across these electrodes were calculated. Difference scores were calculated using the extracted mean amplitudes by subtracting pre-from post-stimulation amplitudes for VR+tACS and VR+Sham. This provided scores indicating change in amplitude within sessions for VR+tACS and VR+Sham. Using difference scores, Pearson correlations were then performed comparing REO and REC for VR+tACS and VR+Sham with EEG ToM and NToM task accuracy and response times for VR+tACS and VR+Sham.

#### 2.9.4 tACS Intensity, Alertness and Blinding

Chi square goodness of fit analyses was used to compare participant-rated blinding for active and sham stimulation. A paired-samples t-test was used to compare tACS stimulation intensity between active and sham stimulation. Finally, alertness for active versus sham and pre versus post stimulation was compared using a repeated measures ANOVA. The researcher remained blinded until all data analysis was completed.

#### 2.9.5 VR Sickness and Immersion

A paired-samples t-test was used to compare VRSQ and PQ sub-scores and the total scores between active and sham conditions.

Assumptions of normality and variance were tested for all analyses. Where assumptions were violated, non-parametric tests were used and reported in the results section. All non-significant findings were subject to Bayes Factor Analyses (BF_10_).

## 3. Results

One participant did not attend their second stimulation session in which they would have received active stimulation. Therefore, the data analysed below is from 14 active sessions and 15 sham sessions.

### 3.1 Behavioural Results

#### 3.1.1 Task Response Time and Accuracy

##### 3.1.1.1 Theory of Mind Task

In the within-session analysis, ToM task response time significantly improved post-active VR-tACS compared to pre-active VR-tACS (*t*(13) = 3.807, *p* = 0.002) as well as post-sham VR-tACS compared to pre- (*t*(14) = 2.818, *p* = 0.014; see **Table 3** and **Figure 5**). There was no significant difference in ToM accuracy post-active VR-tACS (Wilcoxon signed rank test; *z* = −1.067, *p* = 0.298, BF_10_ = 0.408) or post-sham VR-tACS (*t*(14) = −1.133, *p* = 0.276, BF_10_ = 0.453) compared to pre-. On the between session comparisons (i.e., active vs sham sessions), using difference scores between active and sham, there was no significant difference on response time (*t*(13) = 9.136, *p* = 0.894, BF_10_ = 0.272) or response accuracy (*t*(13) = 0.212, *p* = 0.836, BF_10_ = 275) between stimulation conditions (see **Table 3**).

**Figure 5:**
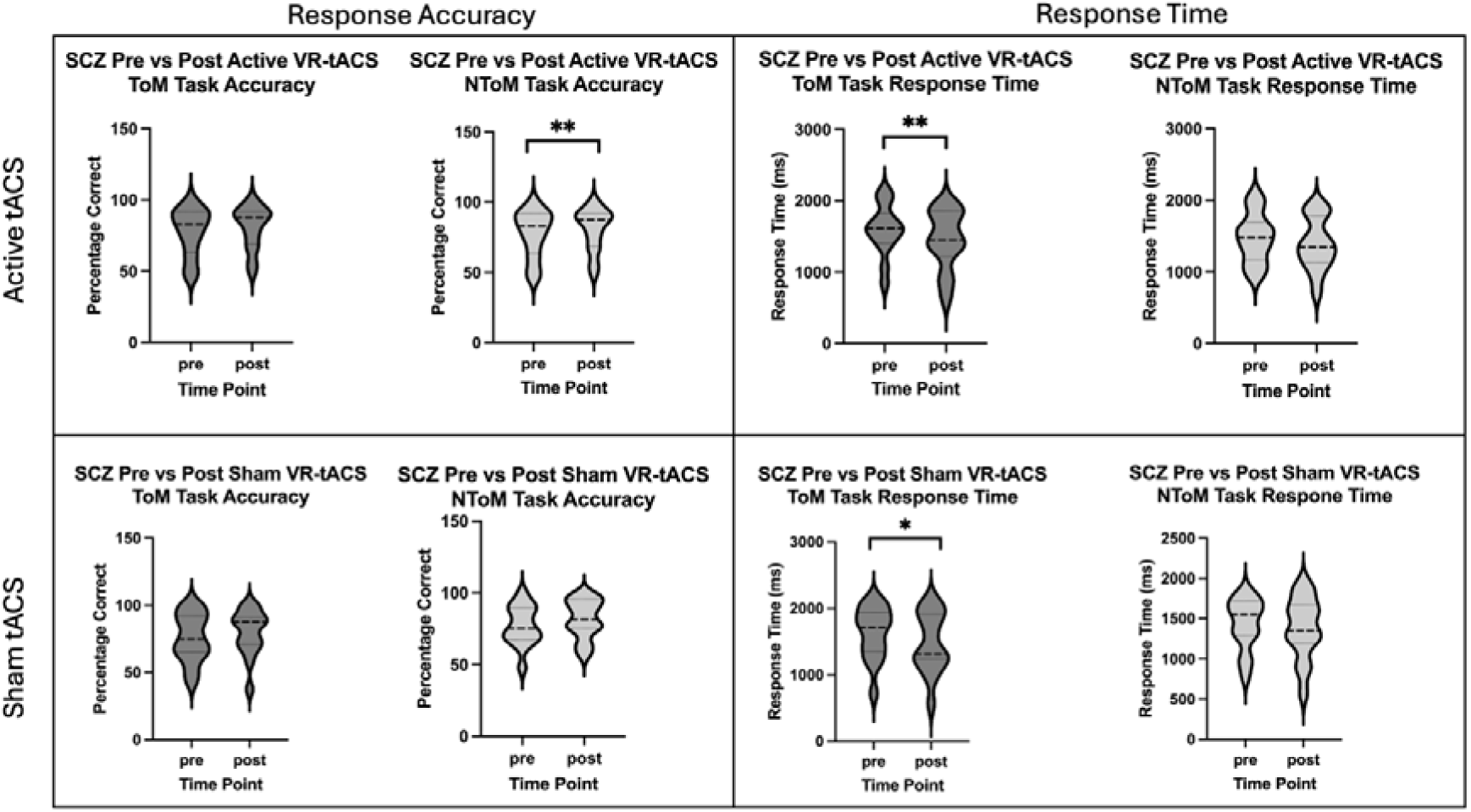
Comparison of response time and accuracy in ToM and NToM tasks pre vs post active and sham tACS combined with VR social cognition training. *p = <0.05, **p=<0.01

**Table 3:** ToM task response time and accuracy.

|  | Accuracy |  |  | Response Time |  |  |
| --- | --- | --- | --- | --- | --- | --- |
|  | Pre | Post | Difference | Pre | Post | Difference |
|  | M(SD) | M(SD) | MD<sup>a</sup>(SD)<sup>b</sup> | M(SD) | M(SD) | MD<sup>a</sup>(SD)<sup>b</sup> |
| Active | 77.446 | 80.911 | 3.466 | 1597.137 | 1451.544 | -145.593 |
| (N=14) | (16.869) | (15.040) | (13.162) | (356.187) | (413.954) | (143.088) |
| Sham | 75.647 | 79.611 | 3.964 | 1632.595 | 1480.174 | -152.421 |
| (N=15) | (17.899) | (16.992) | (13.551) | (395.464) | (454.981) | (209.450) |

<sup>a</sup>mean difference, <sup>b</sup>standard deviation of mean difference

##### 3.1.1.2 Non-theory of mind task

In the within-session analysis, NToM task accuracy significantly improved post-active VR-tACS compared to pre- (*t*(13) = −3.180, *p* = 0.005) but did not improve post-sham VR-tACS compared to pre- (*t*(14) = −1.496, *p* = 0.157, BF_10_ = 0.660; see **Table 4** and **Figure 5**). There was no significant difference in NToM task response time post-active VR-tACS (*t*(13) = 1.449, *p* = 0.171, BF_10_ = 0.638) nor post-sham VR-tACS (*t*(14) = 1.278, *p* = 0.222, BF_10_ = 0.521) compared to pre-. On the between session comparisons (i.e., active vs sham sessions), using difference scores between active and sham, there was no significant difference in response accuracy (*t*(13) = −0.344, *p* = 0.737, BF_10_ = 0.284) or response time (*t*(13) = 1.075, *p* = 0.302, BF_10_ = 0.440) between stimulation conditions (see **Table 4**).

**Table 4:**
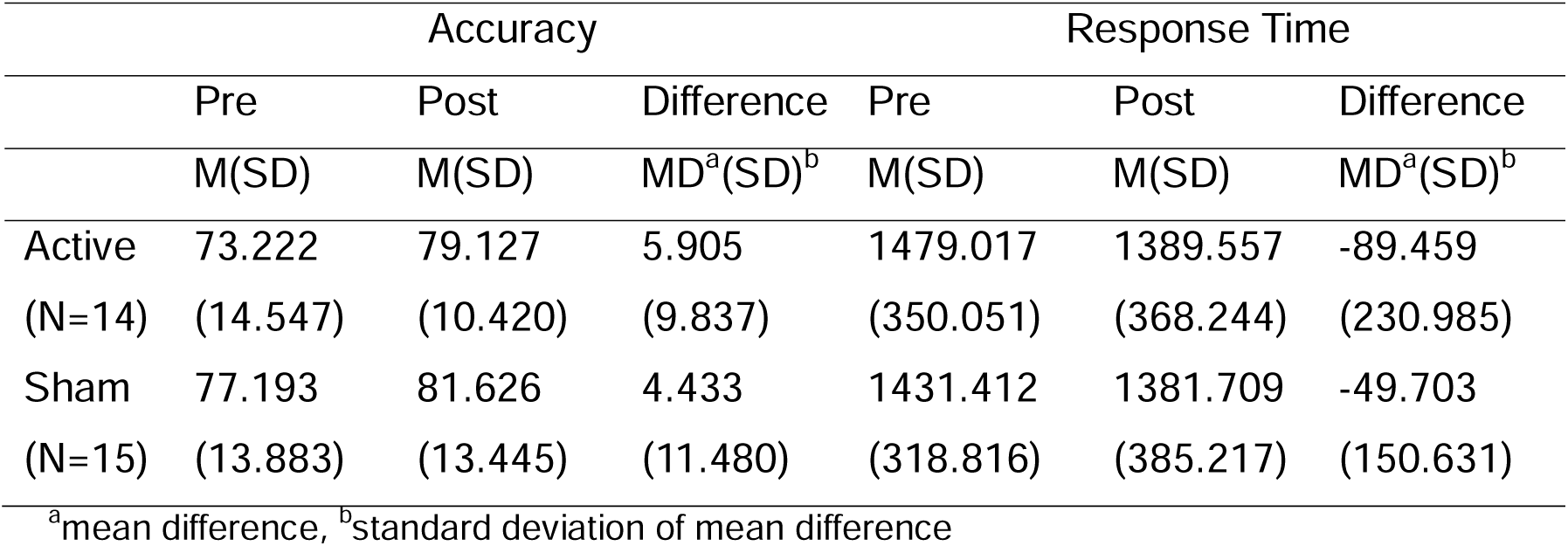
NToM task response time and accuracy.

|  | Accuracy |  |  | Response Time |  |  |
| --- | --- | --- | --- | --- | --- | --- |
|  | Pre | Post | Difference | Pre | Post | Difference |
|  | M(SD) | M(SD) | MD<sup>a</sup>(SD)<sup>b</sup> | M(SD) | M(SD) | MD<sup>a</sup>(SD)<sup>b</sup> |
| Active | 73.222 | 79.127 | 5.905 | 1479.017 | 1389.557 | -89.459 |
| (N=14) | (14.547) | (10.420) | (9.837) | (350.051) | (368.244) | (230.985) |
| Sham | 77.193 | 81.626 | 4.433 | 1431.412 | 1381.709 | -49.703 |
| (N=15) | (13.883) | (13.445) | (11.480) | (318.816) | (385.217) | (150.631) |

<sup>a</sup>mean difference, <sup>b</sup>standard deviation of mean difference

### 3.2 EEG Analysis

#### 3.2.1 Resting State EEG Theta Power Cluster Analyses

##### 3.2.1.1 Resting Eyes Open

There was a significant increase in theta power following VR+tACS across all scalp electrodes compared to baseline (*p* = 0.0016; **Figure 6A**). In contrast, there was no change in theta power following VR+Sham. There was also no significant change in theta power when comparing stimulation conditions (i.e., post VR+tACS vs post VR+Sham).

**Figure 6:**
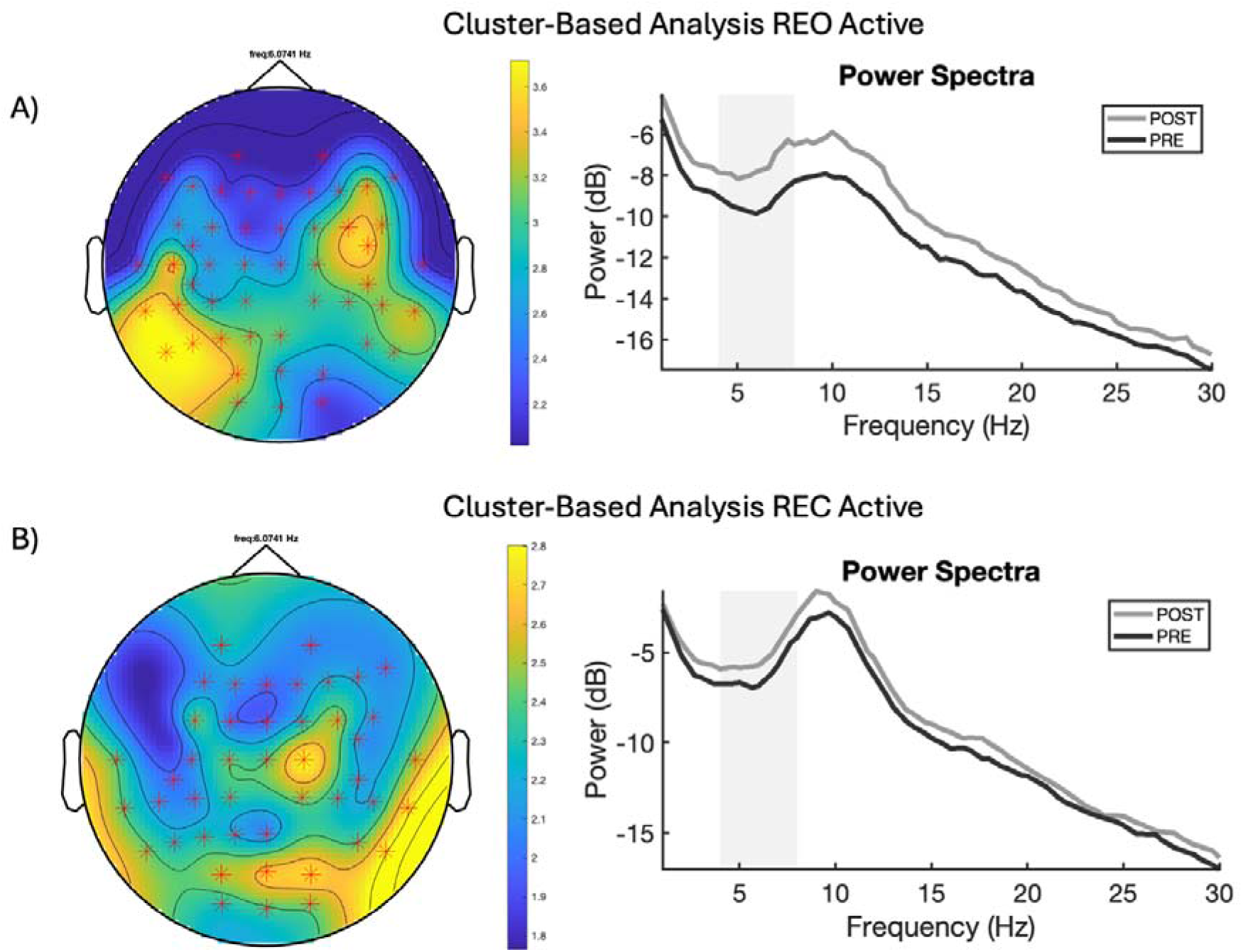
Resting-state theta analysis findings for resting eyes open and eyes closed. Left: Visual representation of cluster-based analysis results. The topographic maps highlight the difference in theta power between the pre- and post VR-tACS conditions for the eyes-open recordings (A) and eyes-closed recordings (B). Right: The spectral power plots show the average theta power across the significant electrode clusters. The grey band on the spectral power plots represent the theta activity band (4-8Hz). Significance level: * p = < 0.01.

##### 3.2.1.2 Resting Eyes Closed

There was a significant increase in theta power post VR+tACS compared to baseline. Similar to the eyes-open results, the cluster was broadly distributed and included the following electrodes FCC6h, CCP5h, CCP6h, Fz, F3, FC1, C3, T7, CP5, CP1, Pz, P3, P7, O1, Oz, O2, P8, CP6, CP2, Cz, T8, FC6, FC2, F4, F8, AF3, F1, FC3, C1, TP7, CP3, P1, P5, PO3, POz, PO4, P6, CP4, TP8, C2, FC4, F6, AF4, F2, FCz (*p* = 0.0099, **Figure 6B**). There was no significant difference in theta power post VR+Sham compared to baseline. There was also no significant difference in theta power when comparing stimulation conditions (i.e., post VR+tACS vs post VR+Sham).

#### 3.2.2 Resting State Theta ROI Analysis

##### 3.2.2.1 Pre vs Post VR-tACS

Due to a violation of the assumption of normality, Wilcoxon signed rank tests were performed for some analyses. There was a significant increase in theta power at rest at the ROI (rTPJ = ‘CP4’; ‘CP6’; ‘TP8’; ‘P6’; ‘P8’) with eyes open post active VR-tACS (*t*(13) = −3.227, *p* = 0.007) and sham (*z* = −2.101, *p* = 0.035) compared to pre- (**Figure 7**). There was a significant increase in theta power at rest at the ROI with eyes closed post-active VR-tACS (*z* = −2.417, *p* = 0.013) but not sham (*z* = −1.250, *p* = 0.229, BF_10_ = 0.335) compared to pre-.

**Figure 7:**
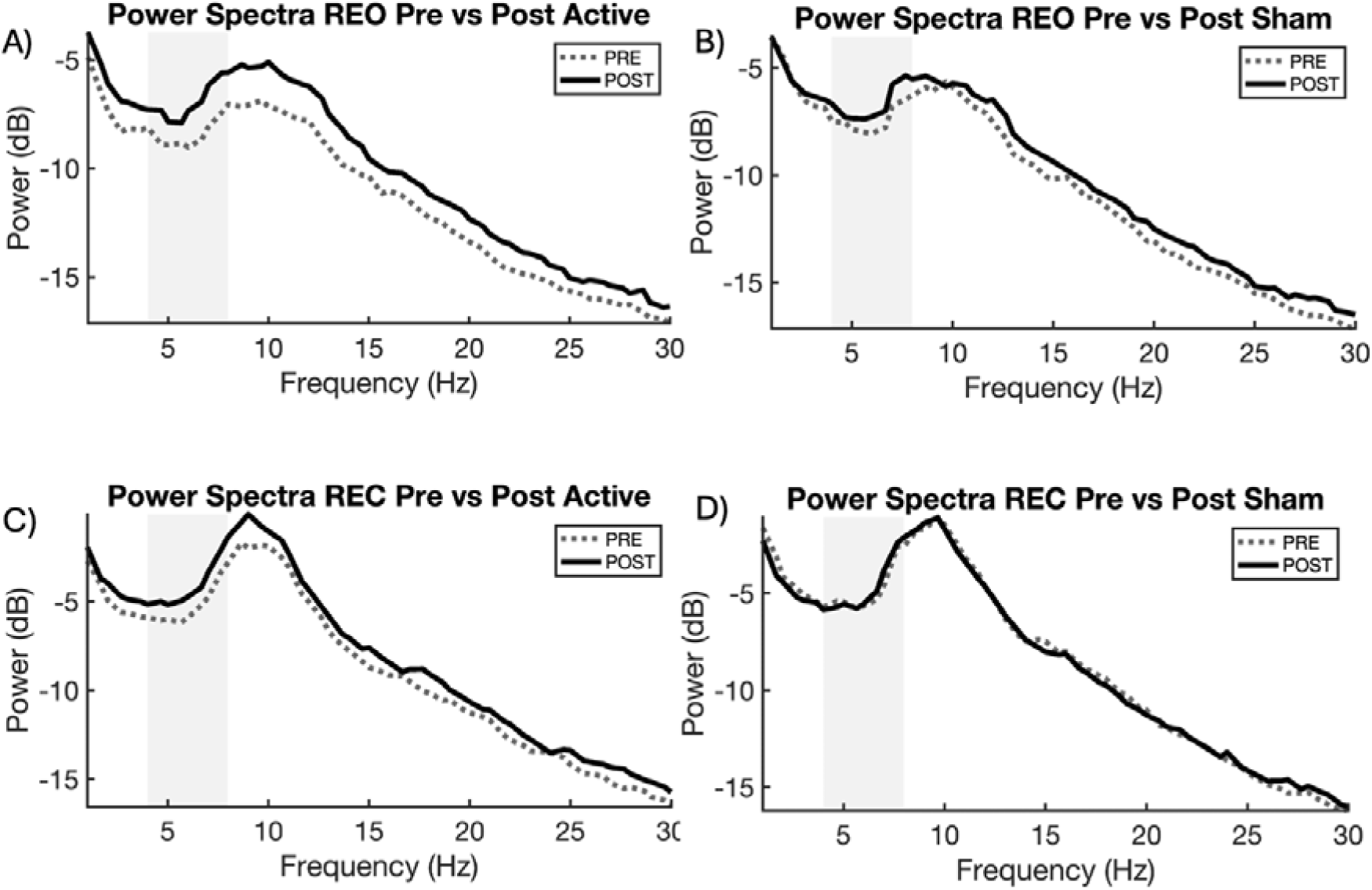
Resting state ROI power spectra plots. Plot A and B show difference in theta power pre vs post VR-tACS for resting eyes open. C and D show differences in theta power pre vs post VR-tACS for resting eyes closed. The grey bar represents theta band activity (4-8Hz).

##### 3.2.2.2 Active vs Sham

Due to a violation of the assumption of normality, Wilcoxon signed rank tests were performed. There was no significant difference in theta power when comparing stimulation conditions for resting eyes open (*z* = 0.220, *p* = 0.855, BF_10_ = 0.520) or resting eyes closed (*z* = −1.475, *p* = 0.153, BF_10_ = 0.575).

#### 3.2.3 Event-Related Potentials

##### 3.2.3.1 TP450

There were no significant differences for ROI or cluster-based statistics. See supplementary materials for additional information.

##### 3.2.3.2 LPC

There were no significant differences for ROI or cluster-based statistics. See supplementary materials for additional information.

### 3.3 Correlations

Exploratory correlations were conducted to investigate the relationship between resting state theta activity findings and behavioural task performance. The data included two outliers (values more than 2SD from the mean). However, these were retained as they were inspected for accuracy and deemed to reflect the true scores. With the outliers retained, there was a significant moderate positive correlation between change in REO theta power and change in ToM task accuracy (*r* = 0.689, *p* = 0.006, see supplementary materials). With outliers removed, the correlation was no longer statistically significant, (*r* = 0.143, *p* = 0.641, BF_10_ = 9.566).

### 3.4 tACS Blinding, Intensity and Alertness

Participants were able to accurately guess better than chance whether they were receiving active tACS (X^2^(1,14) = 4.571, *p* = 0.033) with a moderate degree of confidence in their guess (M = 6.143, SD = 3.570) and sham tACS (X^2^(1,15) = 5.40, *p* = 0.020) with a moderate degree of confidence in their guess (M = 6.429, SD = 2.503). Participants rated tACS intensity as significantly higher in the active condition compared to sham (*t*(13) = −2.160, *p* = 0.050). A repeated measures ANOVA was conducted to investigate differences in participants’ perceived alertness. There was no significant interaction effect of alertness for active and sham tACS (F(1,13) = 0.553, *p* = 0.470).

### 3.5 VR Sickness and Presence

There were no significant differences on ratings of oculomotor VR sickness symptoms (t = 0.922, df = 13, *p* = 0.686) or disorientation symptoms (t = −0.414, df = 13, *p* = 0.686) between active and sham tACS. Additionally, there was no significant difference in overall VR sickness scores (t = 0.540, df = 13, *p* = 0.599). There was no significant difference in how participants experienced presence or immersion in VR between active and sham conditions on any of the PQ sub-scores. See supplementary materials for further information.

## 4. Discussion

This study is the first to investigate the impact of VR social cognition training with concurrent theta tACS on neurophysiological and behavioural measures in people with a diagnosis of schizophrenia or schizoaffective disorder. There was a significant improvement in ToM task response time after VR+tACS and VR+Sham but no significant changes in response time on NToM tasks. NToM task response accuracy improved significantly only after VR+tACS. There was no improvement in response accuracy on the ToM task regardless of stimulation condition. There were no significant differences *between* active and sham VR-tACS on any of the behavioural tasks.

Analysis of the resting-state EEG recordings revealed a significant increase in resting state theta power for both the eyes open and eyes closed conditions following VR+tACS but not VR+Sham. However, there were no significant differences in theta power *between* the two stimulation conditions post-stimulation. Results also suggested a relationship between change in theta power at rest and improvement in ToM task accuracy. However, it did not maintain significance after removal of an extreme outlier. Self-report measures of VR and tACS usability and experience provide support for the feasibility, practicality and efficacy of combining VR with tACS for social cognition in schizophrenia.

### 4.1 ToM and NToM Task Performance

The behavioural task findings show an improvement in ToM task response time following VR and both active and sham tACS. As we used an ‘active’ VR condition for both tACS conditions, these findings suggest the VR social cognitive training may have had an effect on ToM task performance, specifically on response times. Indeed, there is growing evidence that VR social cognition training programs alone can improve social cognitive performance in participants with schizophrenia. For example, Shen et al. (2022) conducted a study comparing three weeks (10 sessions) of VR social cognition and interaction training (VR-SCIT) with traditional SCIT and treatment as usual and found that after VR-SCIT and traditional SCIT participants had higher scores on ToM tasks and improved their emotion perception, hostile attribution skills as well as metacognition. However, it’s also not possible to rule out practice effects, as there is evidence that participants with schizophrenia can experience improved task performance with repetition of the same task (Goldberg, Keefe, Goldman, Robinson, & Harvey, 2010). While care was taken to try to reduce potential confounds and practice effects by counterbalancing tasks and separating task stimuli, participants may still have become familiar with the stimuli and thus, shown improvements in task performance. It would be necessary to include additional stimuli in the ToM computer task in future protocols to reduce the potential for practice effects and to be able to draw more definitive conclusions from findings.

NToM task accuracy was also shown to be improved following VR+tACS but not after VR+Sham. tACS primarily modulates brain oscillations and neural synchronization, which may have a more direct impact on general cognitive functions like working memory and attention – which may be required specifically during the NToM task. Thus, the NToM task may be more sensitive to the effects of tACS than the ToM task. In addition, while we applied theta tACS to the rTPJ to target social cognition, this is a heavily connected brain region and as such stimulation likely indirectly impacted additional brain areas (e.g., prefrontal areas such as the dorsolateral prefrontal cortex related to memory and information processing; Chai, Abd Hamid, & Abdullah, 2018). Finally, the VR task is also engaging neurocognitive processes such as attention, information processing and motor skills that are similarly required for the NToM task. Thus, both the VR and tACS could potentially be engaging neural processes that could improve task performance on the NToM trials and neurocognition more broadly, and to a greater extent than VR alone (i.e., VR+Sham condition).

### 4.2 Resting State EEG

Resting state EEG cluster-based analysis showed an increase in theta power at rest (both eyes open and eyes closed datasets) across the cortex following VR+tACS but not VR+Sham. This finding indicates that VR+tACS was able to successfully alter theta activity in this cohort. Specifically, the results revealed a broad increase in theta power, likely due to the addition of tACS modulating brain activity. To our knowledge, this provides the first evidence that a combined VR-tACS social cognition training protocol can successfully modulate social cognition-related brain activity in schizophrenia.

The finding also demonstrates that quantifiable changes in neural activity can be achieved after stimulation targeting a key region involved in social cognition (i.e., the rTPJ) is combined with VR training. Compared to traditional social cognitive training methods, this has promising potential for reducing intervention times. Indeed, traditional methods have shown brain activity changes after training programs of four weeks, five times a week and up to two years of weekly training (Campos et al., 2016). While the current study did not show improvements in specific behavioural changes alongside neurophysiological changes, other VR-NIBS studies in other fields (e.g., cerebral palsy) have shown these effects after multiple sessions (Collange Grecco et al., 2015). Thus, expansion of the protocol to include multiple sessions may further improve potential for VR-tACS effects on both brain activity and behaviour.

The increase in theta power at rest after VR and active theta tACS compared to baseline, was also widespread across the cortex, rather than being solely confined to the rTPJ stimulation target. While the rTPJ is often considered the hub of ToM activity (Masina et al., 2022; Wang, Callaghan, Gooding-Williams, McAllister, & Kessler, 2016) it is also connected to other networks and cortical regions such as the default mode network (DMN) that includes prefrontal (i.e., medial prefrontal cortex) and parietal (e.g., precuneus and posterior cingulate cortex) areas also involved in social cognitive processing and ToM (Maliske & Kanske, 2022; Menon, 2011; Van Overwalle, 2009). These different networks work together in a complex interplay of feedback and feedforward information to allow ToM processing (Maliske & Kanske, 2022). In schizophrenia, activity within these networks, including theta oscillations, is often diminished (Convertino, Bush, Zheng, Adams, & Burgess, 2022; Hoy et al., 2021; Lee et al., 2011). It is therefore possible that the addition of tACS to the VR protocol may have induced widespread modulation of theta activity by engaging the rTPJ and its connected neural networks. Individually, VR social cognition training has been shown to improve frontoparietal connectivity in the alpha and theta band for people with Autism (De Luca et al., 2021) and tACS has also demonstrated an ability to improve long-range neural connectivity (Helfrich et al., 2014). Thus, it is possible that this combination can also induce connectivity changes, and the findings provide an impetus to explore this with direct measures of connectivity.

When examining the rTPJ ROI on its own, in addition to the findings above, there was also an increase in theta activity at rest with eyes open after VR and sham tACS. As previously noted, VR training alone has been shown to increase theta activity (De Luca et al., 2021) and further research with larger samples would be important to distinguish between the findings.

Finally, a Pearson’s correlation was conducted between change in REO theta brain activity and improvement in ToM task accuracy after VR and active theta tACS. This resulted in moderate positive correlation, confirming a relationship between these two variables. However, this finding should be interpreted with caution as the outliers impacted significance.

### 4.3 Event-Related Potentials

Our hypothesis that active VR-tACS would increase TP450 and LPC ERP peak amplitude during ToM behavioural tasks was not substantiated. While VR and active theta tACS induced a change in theta activity at rest, it may be that the addition of theta tACS did not have the capacity to increase the neural activity threshold enough to see a neurophysiological change in ToM processes during behavioural task performance. In schizophrenia, brain activity during task performance has shown a trend towards being diminished compared to healthy controls (Hoy et al., 2021) and to increase theta activity above the threshold of standard functioning, may require further enhancement. It would be of interest to conduct further research to investigate whether multiple sessions of tACS added to the VR protocol could increase this threshold over time to elicit neural changes while completing the ToM behavioural tasks and therefore, subsequently improve task performance and ToM processing.

### 4.4 VR and tACS Integration

Participants reported tACS stimulation intensity as being significantly higher for active stimulation compared to sham. Participants were also able to successfully guess with certainty greater than chance, whether they were receiving active or sham stimulation. However, there was no significant difference between active and sham stimulation on alertness over time. Finally, VR side effects were minimal, and ratings of presence were moderate.

The current sample of schizophrenia participants rated stimulation intensity higher in the active condition. However, mean intensity ranged from 3-4 out of 10 for both active and sham tACS. In a study by our lab using the same intensity on healthy participants, there was no significant difference for intensity (Gainsford et al., 2025a). This may indicate an increased level of scalp sensitivity for the clinical group. Indeed, research shows that people with schizophrenia can experience lower pain thresholds (Sakson-Obada, 2024). It is important to take this into consideration in future protocols and investigate the effects of lower stimulation intensities over more sessions to determine if this provides additional comfort with clinical benefits.

Overall, these findings provide evidence that participants had few difficulties with carrying out the required tasks during the session while wearing VR, tACS and an EEG cap at the same time as navigating the physical and virtual space and receiving tACS. They were able to feel immersed in the virtual environment to a moderate degree and experienced minimal VR and tACS side effects. In a population where social cognitive and cognitive difficulties are common and can adversely impact task performance (Gainsford et al., 2025b; Green et al., 2019), this is a promising result for implementing combined VR and tACS protocols for future interventions.

## 5. Limitations and Future Directions

While VR+tACS showed a significant increase in theta activity at rest, there was no specific differentiation between VR+tACS and VR+Sham on ToM behavioural task performance. There was, however, a significant improvement in NToM task accuracy during VR+tACS compared to VR+Sham. This indicates that the combination of VR+tACS could produce some change for cognition and potentially a different task or protocol with more sensitive ToM stimuli could show social cognitive differences, specifically. There was also no specific differentiation in ERPs during behavioural tasks. This indicates there may be a need for increasing the number of intervention sessions to determine if there could be an increase above theta activation threshold to ToM task performance (and thereby ToM processing) over time. While the sample size is reasonable for a pilot study, the research may have been underpowered and there would also be benefit from conducting future studies with a larger sample size. Additionally, because VR training itself is able to elicit brain changes, it would be helpful in future research to include a “true sham” condition where a control VR protocol is combined with sham tACS. This would help to provide further evidence for the efficacy of concurrent VR-tACS as an intervention. tACS blinding was not effective for this study, with the schizophrenia sample finding the active tACS condition intensity significantly higher than sham. Future research investigating varied stimulation intensities and piloting the intensities with clinical participants would be of benefit. Finally, the current study uses individual measures of tACS and VR usability and experience to provide support for the feasibility and practicability of the application of the concurrent VR-tACS protocol. However, as more VR-NIBS protocols are trialled for treatment, it would be important to develop user experience measures that are specific to combined VR-tACS protocols. This would allow further refinement of these types of interventions and create more user-friendly, immersive and effective protocols.

## 6. Conclusion

The current study provides evidence of the potential for the addition of tACS to VR social cognition training to modulate brain activity and behavioural task performance in schizophrenia, thus, supporting the feasibility and efficacy of a combined VR-tACS protocol for social cognition in schizophrenia. The protocol allowed the integration of VR, tACS and EEG measures and provides the foundation to continue development of concurrent VR social cognition training and tACS in larger scale, multi-session trials as well as with more clinical populations.

## Data Availability

All data produced in the present study are available upon reasonable request to the authors

## Acknowledgements

We gratefully acknowledge the time and involvement from the participants. This research would not have been possible without them.

