## Supplementary Data for "Concurrent virtual reality and transcranial alternating current stimulation for social cognition and neural activity in schizophrenia: A proof-of-concept study"

1. Analysis

1.1 Resting State Analysis Epochs

**Table S1**: *Mean number of epochs in resting state data analysis*

| | | Active (N=14) | Sham (N=15) |
|---|---|---|---|
| Condition | Time Point | M(SD) | M(SD) |
| REO | Pre | 56.36(3.10) | 59.00(4.38) |
| | Post | 56.79(1.63) | 57.07(2.37)) |
| REC | Pre | 56.71(2.16) | 58.00(2.75) |
| | Post | 57.14(1.83) | 57.80(2.57) |

REO = resting eyes open, REC = resting eyes closed

1.2 ERP Analysis Epochs

**Table S2**: *Mean number of epochs in behavioural task data*

| | | Active (N=14) | Sham (N=15) |
|---|---|---|---|
| Condition | Time Point | M(SD) | M(SD) |
| ToM | Pre | 23.71(0.47) | 23.07(1.03) |
| | Post | 23.50(0.86) | 23.67(0.72) |
| NToM | Pre | 46.43(2.10) | 47.00(1.77) |
| | Post | 46.21(1.89) | 45.80(2.27) |

ToM = theory of mind task, NToM = non-theory of mind task

2. Results

2.1 Event-Related Potentials

2.1.1 TP450

There were no significant differences in theta power for the TP450 ERP at the CP6 ROI during the ToM task when comparing post VR and active tACS with pre ($t(13) = 0.603$, $p = 0.557$) or sham ($t(14) = -1.307$, $p = 0.212$). There were no significant differences in theta power for the TP450 ERP at the CP6 ROI during the NToM task when comparing post VR and active tACS with pre ($t(13) = -0.998$, $p = 0.337$) or sham ($t(14) = -0.093$, $p = 0.927$). There were no significant differences in theta power between VR and active and sham tACS sessions during the ToM task ($t(13) = 0.919$, $p = 0.375$) or the NToM task (Wilcoxon $z = 0.847$, $p = 0.426$).

There were no significant differences in theta power for the TP450 ERP at the rTPJ ROI (CP4, CP6, TP8, P6, P8) during the ToM task post VR and active tACS ($t(13) = 0.342$, $p = 0.738$) or sham ($t(14) = -1.044$, $p = 0.314$). There were no significant differences in theta power for the TP450 ERP at the rTPJ ROI (CP4, CP6, TP8, P6, P8) during the NToM task post VR and active tACS ($t(13) = -1.315$, $p = 0.211$) or sham ($t(14) = -0.240$, $p = 0.814$). There were no significant differences in theta power between VR and active and sham tACS sessions during the ToM task ($t(13) = 0.635$, $p = 0.536$) or the NToM task ($t(13) = -1.055$, $p = 0.310$).

2.1.2 LPC

There were no significant differences in theta power for the LPC ERP at the Pz ROI during the ToM task when comparing post VR and active tACS with pre ($t(13) = -0.426$, $p = 0.677$) or sham ($t(14) = -1.929$, $p = 0.074$). There were no significant differences in theta power for the LPC ERP at the Pz ROI during the NToM task when comparing post VR and active tACS with pre ($t(13) = 0.373$, $p = 0.715$) or sham ($t(14) = 2.051$, $p = 0.059$). There were no significant differences in theta power between VR and active and sham tACS sessions during the ToM task ($t(13) = 1.321$, $p = 0.209$) or the NToM task ($t(14) = -0.613$, $p = 0.550$).

There were also no significant differences in theta power for the LPC ERP at the Pz, Cz ROI during the ToM task post VR and active tACS ($t(13) = -0.490$, $p = 0.632$). The result for post VR and sham tACS is reported in the main paper. There were also no significant differences in theta power for the LPC ERP at the Pz, Cz ROI during the NToM task post VR and active tACS ($t(13) = 0.647$, $p = 0.529$) or sham ($t(14) = 1.607$, $p = 0.130$). There were no significant differences in theta power between VR and active and sham tACS sessions during the ToM task ($t(13) = 1.057$, $p = 0.310$) or the NToM task ($t(13) = -0.265$, $p = 0.795$).

### 2.2 Correlation

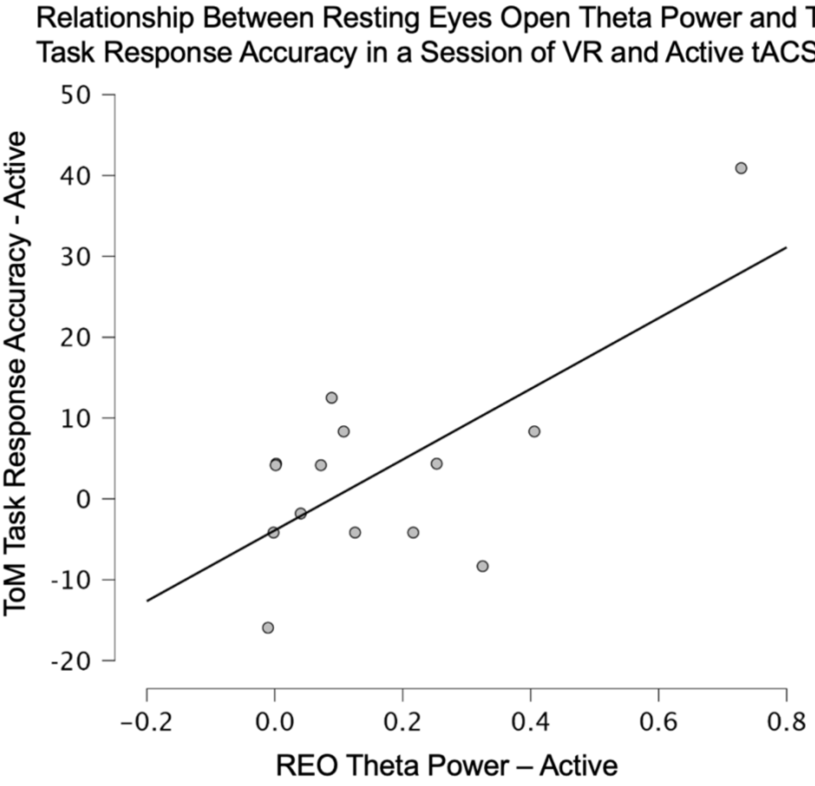


**Figure S1**: *Relationship between difference in resting eyes open theta power (pre vs post) VR and concurrent active theta tACS and difference in ToM task response accuracy (pre vs post) VR and concurrent active theta tACS.*

### 2.3 VR Side Effects and Presence

The presence questionnaire results showed no significant difference between active and sham sessions on any of the sub-scores. These include realism ($t(13) = 2.113$, $p = 0.054$), possibility to act ($t(13) = -0.069$, $p = 0.946$), quality of interface ($t(13) = -0.874$, $p = 0.398$), possibility to examine ($t(13) = 0.492$, $p = 0.631$), self-evaluation ($t(13) = 1.412$, $p = 0.181$) and sound ($t(13) = -0.658$, $p = 0.522$).

**Table S3**: *Descriptive statistics of VR measures*

| | | Active (N=14) | | Sham (N=15) | |
|---|---|---|---|---|---|
| Measure | Sub-score | M(SD) | Range | M(SD) | Range |
| VR Sickness Questionnaire | Oculomotor | 17.857 (17.559) | 0-50 | 20.556 (16.627) | 0-50 |
| | Disorientation | 13.333 (12.541) | 0-40 | 11.556 (12.464) | 0-33.33 |
| | Total | 15.595 (13.671) | 0-38.33 | 16.056 (14.105) | 0-41.67 |
| Presence Questionnaire | Realism | 32.071 (11.418) | 12-46 | 35.933 (8.556) | 14-47 |
| | Possibility to Act | 20.786 (5.177) | 10-27 | 20.60 (3.888) | 14-25 |
| | Quality of Interface | 14.357 (3.734) | 6-20 | 13.60 (3.869) | 8-19 |
| | Possibility to Examine | 15.50 (4.220) | 6-21 | 16.067 (3.494) | 8-21 |
| | Self-evaluation | 10.857 (3.035) | 3-14 | 11.667 (1.759) | 8-14 |
| | Sounds | 16.571 (3.031) | 12-21 | 15.933 (3.348) | 7-21 |
